# A unique SARS-CoV-2 spike protein P681H strain detected in Israel

**DOI:** 10.1101/2021.03.25.21253908

**Authors:** Neta S. Zuckerman, Shay Fleishon, Efrat Bucris, Dana Bar-Ilan, Michal Linial, Itay Bar-Or, Victoria Indenbaum, Merav Weil, Israel National Consortium for SARS-CoV-2 sequencing, Ella Mendelson, Michal Mandelboim, Orna Mor

## Abstract

Routine detection, surveillance and reporting of SARS-CoV-2 novel variants is important, as these threaten to hinder vaccination efforts. Herein we report a local novel strain that includes a non-synonymous mutation in the spike (S) protein - P681H and additional synonymous mutations. The P681H Israeli strain has not been associated with higher infection rates and was neutralized by sera from vaccinated individuals in comparable levels to the B.1.1.7 strain and a non-P681H strain from Israel.

## Introduction

The severe acute respiratory syndrome coronavirus 2 (SARS-CoV-2) outbreak is in the midst of its third wave in Israel, with increasing efforts to sequence complete SARS-CoV-2 genomes to detect known and emerging variants. Herein we report a local novel strain that includes a non-synonymous mutation in the spike (S) protein - P681H and additional synonymous mutations. The P681H Israeli strain has not been associated with higher infection rates and was neutralized by sera from vaccinated individuals in comparable levels to the B.1.1.7 strain and a non-P681H strain from Israel.

## Results

### Characterization of the Israeli P681H strain

The P681H strain presents as a cluster in a phylogenetic tree of all sequenced samples in Israel, distinct from the B.1.1.7 Pangolin lineage, which also contains the P681H mutation (Figure 1A). The strain originates from the B.1.1.50 Pangolin lineage, in which the majority of the sequences are from Israel (70%), Palestine (12%) and the UK (12%) [1]. It is characterized by a non-synonymous mutation in the S protein - P681H (C23604A) and additional four synonymous mutations: Nsp3:C7765T, Nsp12b:C13821T, Nsp16:T21111C and C29545A. The strain carrying these mutations is unique to Israel, aside for 2 sequences originating in Palestine. A sub-clade of the strain contains an additional non-synonymous mutation in the S protein - A27S (G21641T) (Figure 1B).

**Figure 1.**
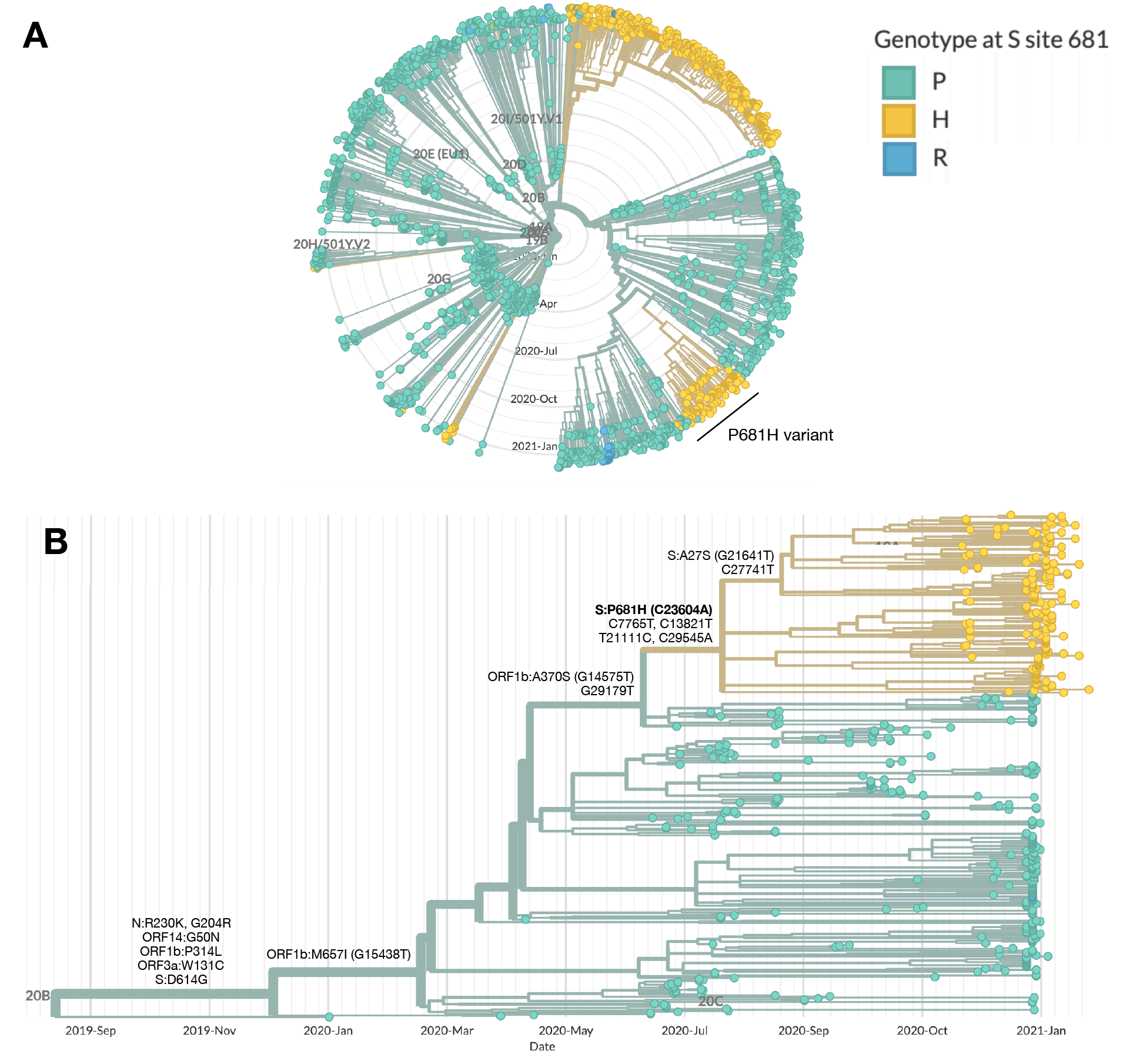
Characterization of P681H strain in Israel within the B.1.1.50 lineage. Phylogenetic trees highlighting the genotype at site 681 in the S protein, with the wild-type proline (P) in green and the mutations histidine (H) and arginine (R) in yellow and blue, respectively. **(A)** SARS-CoV-2 whole genomes sequenced in Israel from March 2020 to January 2021 (n=2482). The main clusters harboring the P681H mutation are the B.1.1.7 (20I/501Y.V1) and the P681H variant. **(B)** SARS-CoV-2 whole genomes of the B.1.1.50 lineage (n=489). The P681H cluster is composed of local viruses only, and of 2 sequences from isolates identified in Palestine, and includes a sub-cluster with an additional S protein non-synonymous mutation, A27S. Phylogenetic trees were created with Nextstrain Augur pipeline and visualized with Auspice [2]. Non-Israeli B.1.1.50 lineage sequences were identified with using Pangolin [1] and downloaded from GISAID.

Until January 2021, an overall of 181 individuals were infected with the P681H strain. These were mostly randomly sampled from districts from all over Israel, with no gender or age bias (Table 1).

**Table 1.** Epidemiology of P681H Israeli variant. Frequency and patient-related information of the P681H Israeli strain. Ret. abroad is return from abroad; Jud & Sam is Judea and Samaria.

|  |  | November | December | January |
| --- | --- | --- | --- | --- |
| gender | Male | 10 | 30 | 25 |
|  | Female | 8 | 24 | 23 |
|  | Unknown | 9 | 17 | 35 |
| age |  | 39.3 ± 21.3 | 33.8 ± 25.3 | 35.8 ± 22 |
| reason | random | 27 | 55 | 80 |
|  | outbreaks |  | 10 |  |
|  | ret. abroad |  | 6 | 3 |
| district | Northern | 3 | 7 | 14 |
|  | Haifa | 4 | 2 | 5 |
|  | Central | 4 | 19 | 32 |
|  | Tel Aviv | 1 | 11 | 10 |
|  | Jerusalem | 1 | 9 | 9 |
|  | Jud & Sam | 1 |  | 5 |
|  | Southern | 1 | 4 | 3 |
|  | Unknown | 12 | 19 | 5 |
| total |  | 27 | 71 | 83 |

### Identification of P681H mutation in sewage

The P681H mutation was also frequently identified by SARS-CoV-2 whole genome sequencing of samples from waste-water treatment plants in 9 locations across Israel, that were collected once a month, August 2020 - January 2021 (Figure 1). As the P681H mutation is also found in the B.1.1.7 variant, its frequency has increased in some locations since late December 2020, following the first introduction of B.1.1.7 into Israel. Notably, the mutation was already observed with 5% frequency as early as October 2020 in Rahat (city in the South of Israel) and in November, at a frequency of 98% in Natanya and Haifa, located at the center and north of Israel, respectively (Figure 2).

**Figure 2.**
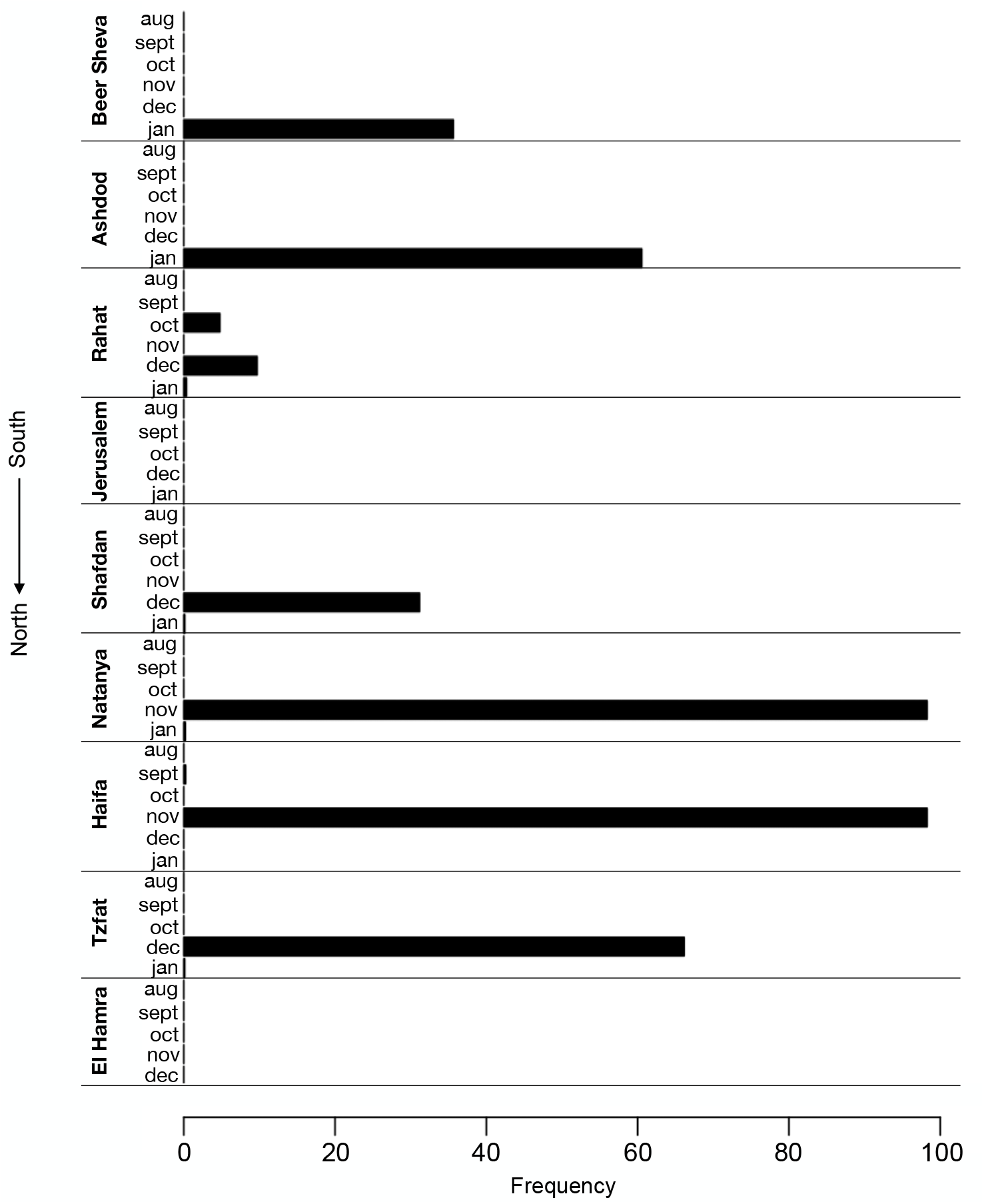
Identification of the P681H mutation in sewers across Israel. Frequency of P681H mutation in SARS-CoV-2 genomes sequenced from 9 wastewater treatment plants across Israel each month in August 2020 – January 2021. The frequency of the P681H mutation in each region was estimated by measuring the fraction of the mutation from the total number of nucleotides mapped to the position (i.e., depth of sequencing).

### Effective neutralization of the P681H strain

The neutralization potency of antibodies against the local P681H strain was compared to the neutralization of other strains circulating in Israel. VERO-E6 cells were infected with (a) Israeli P681H strain (isolate hCoV-19/Israel/CVL-45176-P681H-ngs/2020), (b) non-P681H strain from Israel (isolate hCoV-19/Israel/CVL-45526-ngs/2020) and (c) B.1.1.7 strain (isolate hCoV-19/Israel/CVL-46879-ngs/2020). The different strains were pre-incubated with serial dilutions of serum samples obtained from individuals vaccinated with two doses of the Pfizer vaccine. Results demonstrate comparable neutralization of the sera against the P681H Israeli strain, the B.1.1.7 strain and the non-P681H strain (Figure 3).

**Figure 3.**
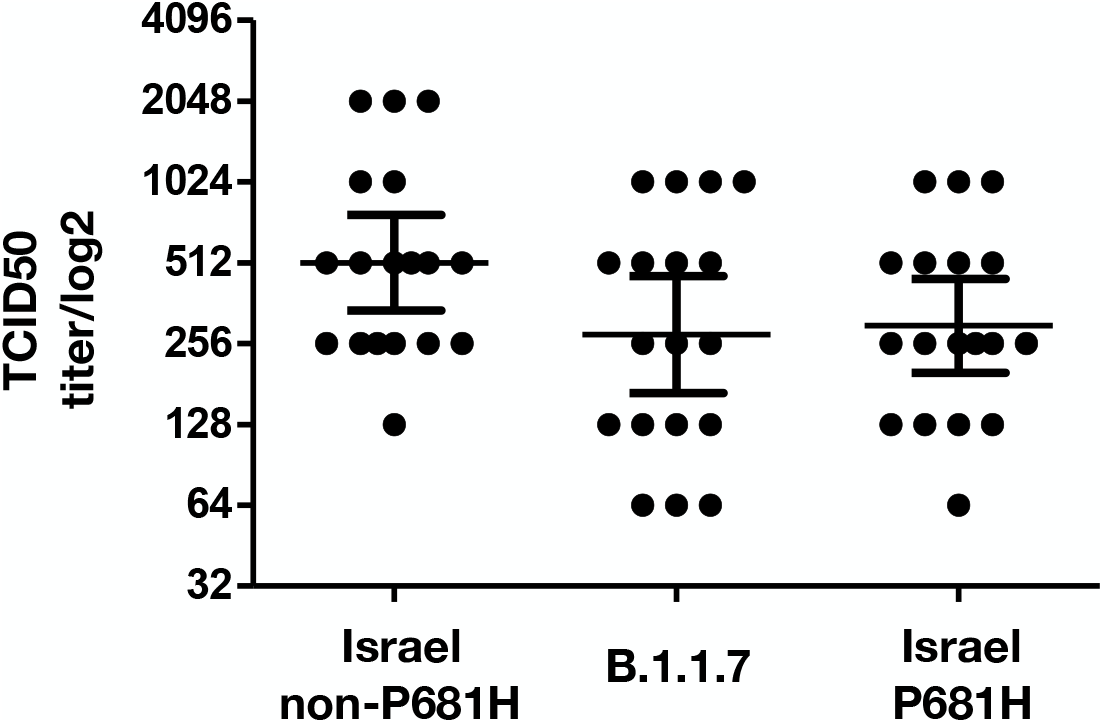
Neutralization of the P681H Israeli strain. Neutralization assays were carried out with VERO-E6 cells infected with the P681H Israeli strain, B.1.1.7 strain and non-P681H strain from Israel, using sera from vaccinated individuals. On day 6, plates were colorized overnight with Gentian violet + 4% formaldehyde solution for virus neutralization. Titers were calculated by qualitative measurements of the cytopathic effect for each patient. Bars represent the geometric mean titer (GMT) and 95% confidence intervals.

## Discussion and conclusions

The P681H mutation has been observed as early as March 2020 in samples worldwide, e.g. in Nigeria [3] and Hawaii [4] and also characterizes the globally spreading B.1.1.7 variant. Herein, we report 181 sequences that are part of the B.1.1.50 lineage, but form a unique local strain that harbors the P681H mutation along with additional defining mutations. The P681H strain has been identified in clinical samples since November 2020, however the P681H mutation has been identified in Israel already in October 2020 via SARS-CoV-2 wastewater sequencing.

The P681H is located in the vicinity of the furin cleavage site in the S protein. It is postulated to enhance transmissibility of the virus by facilitating a conformational change in the S protein following protease activity at the cell membrane [3]. Previous serum neutralization assays for VSV-based SARS-CoV-2 pseudoviruses expressing the S protein with the P681H mutation demonstrated no significant changes in the titer required for neutrazliation compared to the unmutated S protein [4]. Additionally, no significant changes in the titer were needed for neutralization of the WT SARS-CoV-2 compared to a viral sample from the B.1.1.7 lineage (in which P681H is one of its defining mutations), using sera from convalescent patients or individuals vaccinated with the mRNA-1273 vaccine from Moderna [5]. Similar to these reports, this study demonstrates comparable neutralization of the P681H Israel strain, non-P681H strain and the B.1.1.7 variant by sera from vaccinated individuals.

Overall, as the P681H local strain was neutralized as efficiently as commonly circulating strains, was not associated with escalated infection or spread and given that the P681H mutation is observed in additional SARS-CoV-2 strains worldwide, this emerging local variant is currently not defined as a variant of concern. Nevertheless, it is still monitored by routine next generation sequencing in Israel.

## Methods

### Sample collection for sequencing

Random sampling and collection of SARS-CoV-2 PCR-positive samples for sequencing complete viral genomes is routinely conducted in Israel as part of a national effort for monitoring SARS-CoV-2 variants, starting December 2020.

### Library preparation and sequencing

Viral genomes were extracted from 200µl respiratory samples with the MagNA PURE 96 (Roche, Germany), according to the manufacturer instructions. Libraries were prepared using COVID-seq library preparation kit, as per manufacturer’s instructions (Illumina). Library validation and mean fragment size was determined by TapeStation 4200 via DNA HS D1000 kit (Agilent). Libraries were pooled, denatured and diluted to 10pM and sequenced on NovaSeq (Illumina).

### Bioinformatics analysis

Fastq files were subjected to quality control using FastQC (www.bioinformatics.babraham.ac.uk/ projects/fastqc/) and MultiQC [6] and low-quality sequences were filtered using trimmomatic [7]. Sequences were mapped to the SARS-CoV-2 reference genomes (NC_045512.2) using Burrows-Wheeler aligner (BWA) mem [8]. Resulting BAM files were sorted, indexed and subjected to quality control using SAMtools suite [9]. Coverage and depth of sequencing was calculated from sorted bam files using a custom python script. A consensus sequence was constructed for each sample using iVar (https://andersen-lab.github.io/ivar/html/index.html), where positions with <5 nucleotides were determined as Ns, and converted to a fasta file using seqtk (https://github.com/lh3/seqtk). Sequences were aligned to SARS-CoV-2 reference genome (NC_045512.2) using MAFFT [10].

Mutation calling, translation to amino acid and identification of P681H variant sequences were done in R with a custom code using Bioconductor package Seqinr [11]. Sequences were further analyzed together with additional sequences identified as belonging to the background lineage B.1.1.50 downloaded from GISAID [12].

Phylogenetic trees were constructed using the Augur pipeline [2]. Sequences were aligned to SARS-CoV-2 reference genome (NC_045512.2) using MAFFT [10], and a time-resolved phylogenetic tree was constructed with IQ-Tree [13] and TreeTime [14] under the GTR substitution model and visualized with auspice [2]. Lineage nomenclature was attained from Pangolin Lineages [15].

Analysis of sewage samples was conducted using a custom R code and the Bioconductor package Rsamtools [16]. BAM files were imported to R and the frequency of each mutated position along the genome, out of the total number of nucleotides covering that position, was obtained. The frequency in position 681 in the spike protein was recorded for each sample.

### Neutralization assays

VERO-E6 cells at concentration of 20*103/well were seeded in sterile 96-wells plates with 10% FCS MEM-EAGLE medium, and stored at 37°C for 24 hours. One hundred TCID50 of P681H variant, non-P681H and B.1.1.7 isolates were incubated with inactivated sera diluted 1:10 to 1:1280 in 96 well plates for 60 minutes at 33ºC. Virus-serum mixtures were added to the Vero E-6 cells and incubated for five days at 33ºC, after which Gentain violet staining (1%) was used to stain and fix the cell culture layer. Neutralizing dilution of each serum sample was determined by identifying the well with the highest serum dilution without observable cytopathic effect. A dilution equal to 1:10 or above was considered neutralizing.

## Data Availability

Sequence data has been deposited to GISAID.

## Acknowledgments

The National Covid-19 Information & Knowledge Center.

